# Clinical, sociodemographic, and genetic predictors of depressive episode duration in the UK Biobank

**DOI:** 10.64898/2026.08.19.26360769

**Authors:** Louise S. Schindler, Madhurbain Singh, Emerie Sheridan, Chris Wai Hang Lo, Michelle Kamp, Cathryn M. Lewis

## Abstract

**Background:** The course of major depressive disorder is heterogeneous, with UK Biobank (UKB) participants reporting episode durations ranging from <1 month to >24 months. Here, we identify predictors of episode duration, characterise its genetic architecture, and examine links to treatment seeking and response.

**Methods:** In UKB participants meeting criteria for major depressive disorder, we examined clinical, sociodemographic, and genetic predictors of short (0-3 months) and long (>24 months) episode duration, fitted in predictor-specific, domain-level, and combined models. We also conducted genome-wide association studies in European-ancestry participants (n = 40,858) and estimated common-variant heritability.

**Results:** Clinical features were most informative: higher childhood trauma scores, a stressful trigger, and recurrence showed the most consistent associations with short and long durations across models (OR_combined_: short = 0.75-0.95; long = 1.13-1.45; all p≤0.02). Higher neuroticism scores were also associated with both durations (OR_combined_: short = 0.977; long = 1.053; p<0.001). Polygenic risk for depression was associated with episode duration, though its independent contribution was modest. Long episodes were more predictable than short in validation analyses (AUC = 0.705 vs 0.601) and were associated with greater treatment engagement but lower perceived benefit; SNP-based heritability was nominally significant.

**Conclusions:** Clinical features captured most of the predictable variance in episode duration, with the same predictors largely operating in opposite directions for short and long episodes, consistent with a continuum of chronicity. Those at risk for long episodes emerge as a priority for early identification and intervention.

## Introduction

Major depressive disorder (MDD) is one of the most common and burdensome conditions worldwide (Yan et al., 2024). Individuals differ considerably across symptom profiles, comorbidities, treatment response, and long-term course, reflecting MDD’s substantial heterogeneity, with implications for prognosis and care (Milaneschi et al., 2020; Musliner et al., 2016). However, the duration of depressive episodes has received comparatively little investigation as a factor contributing to depression heterogeneity.

Episode duration follows a right-skewed distribution: many people’s episodes remit within a few months, but a substantial minority have episodes that persist for over a year (Eaton et al., 2008; Patten, 2006). A smaller group has episodes lasting over two years, qualifying for a persistent depressive disorder (PDD) diagnosis (Schramm et al., 2020). In the Netherlands Mental Health Survey and Incidence Study (NEMESIS) and its follow-up, median episode duration was approximately 3-6 months, yet 12-20% of individuals had not recovered at 24-36 months (Spijker et al., 2002; ten Have et al., 2017), a pattern consistent with findings from the Group for Longitudinal Affective Disorders Study (GLADS), a treatment-naïve Japanese cohort (Furukawa et al., 2000). Importantly, this variation is unlikely to reflect differential treatment response alone, as placebo arms of antidepressant trials show markedly heterogeneous response distributions, pointing to individual-level factors (Stone et al., 2022).

Understanding which factors contribute to episode duration is crucial, as longer episodes confer substantial personal, clinical, and societal burdens. Clinically, longer episodes and longer durations of untreated depression are associated with poorer symptomatic and functional recovery and an increased risk of treatment-resistant depression (Ghio et al., 2015; Kautzky et al., 2019). These prolonged episodes place a growing burden on patients, healthcare services, and the wider economy, with healthcare demand rising as depression persists (Pappa et al., 2024). While some studies have examined clinical factors contributing to episode duration (Melartin et al., 2004; ten Have et al., 2017), no study has to our knowledge jointly modelled clinical, sociodemographic, and genetic predictors of depressive episode duration within a single framework. The availability of large biobank cohorts with both deep phenotyping and genetic data makes this integrated approach both feasible and timely.

In this study, we examined predictors of depressive episode duration in UK Biobank (UKB) participants meeting criteria for a major depressive episode, contrasting short (0-3 months) and long (>24 months) episodes. We tested clinical, sociodemographic, and genetic predictors individually and in combination, evaluating predictive performance in a held-out sample; conducted a genome-wide association study (GWAS) and estimated SNP-based heritability; and examined associations with treatment-seeking behaviour and perceived treatment effectiveness. Together, these analyses characterise episode duration as a clinically meaningful phenotype and allow us to examine whether long episodes represent the upper tail of a continuum of chronicity or a distinct subgroup.

## Methods

### Participants

The UKB is a large prospective cohort of about 500,000 UK volunteers, aged 40-70 at recruitment between 2006 and 2010 (Allen et al., 2014). Data collected at baseline include medical history and sociodemographic and lifestyle variables. Alongside these baseline measures, UKB administered two online mental health questionnaires in 2016 (MHQ1; Category 136) and 2022 (MHQ2; Category 1502) (Davis et al., 2025); [https://osf.io/c65t7/overview]. Our analyses focused on participants from MHQ2, which has a wider range of information related to depression.

Participants were asked about MDD symptoms during their worst depressive episode in the Composite International Diagnostic Interview - Short Form (CIDI-SF). We identified participants meeting the criteria for a lifetime depressive episode, endorsing at least one cardinal symptom (persistent sadness or anhedonia), functional impairment in daily life, and a minimum of five other symptoms, including weight change, sleep disturbance, fatigue, feelings of worthlessness, difficulty concentrating, and thoughts of death. Supplementary Information (SI) Section 1 provides UKB field codes and further details.

From these cases, we retained participants who had polygenic scores (PGS) available for the psychiatric and treatment-response phenotypes of interest, were genetically similar to European ancestry (EUR-like; to reduce population stratification in genetic analyses), had no prior or probable diagnosis of schizophrenia, bipolar disorder, or mania, and had responded to the question on worst-episode duration. This yielded a primary analysis sample of 33,139 MHQ2 participants; missing sociodemographic and clinical variables reduced sample sizes further for some models (Figure 1).

**Figure 1.**
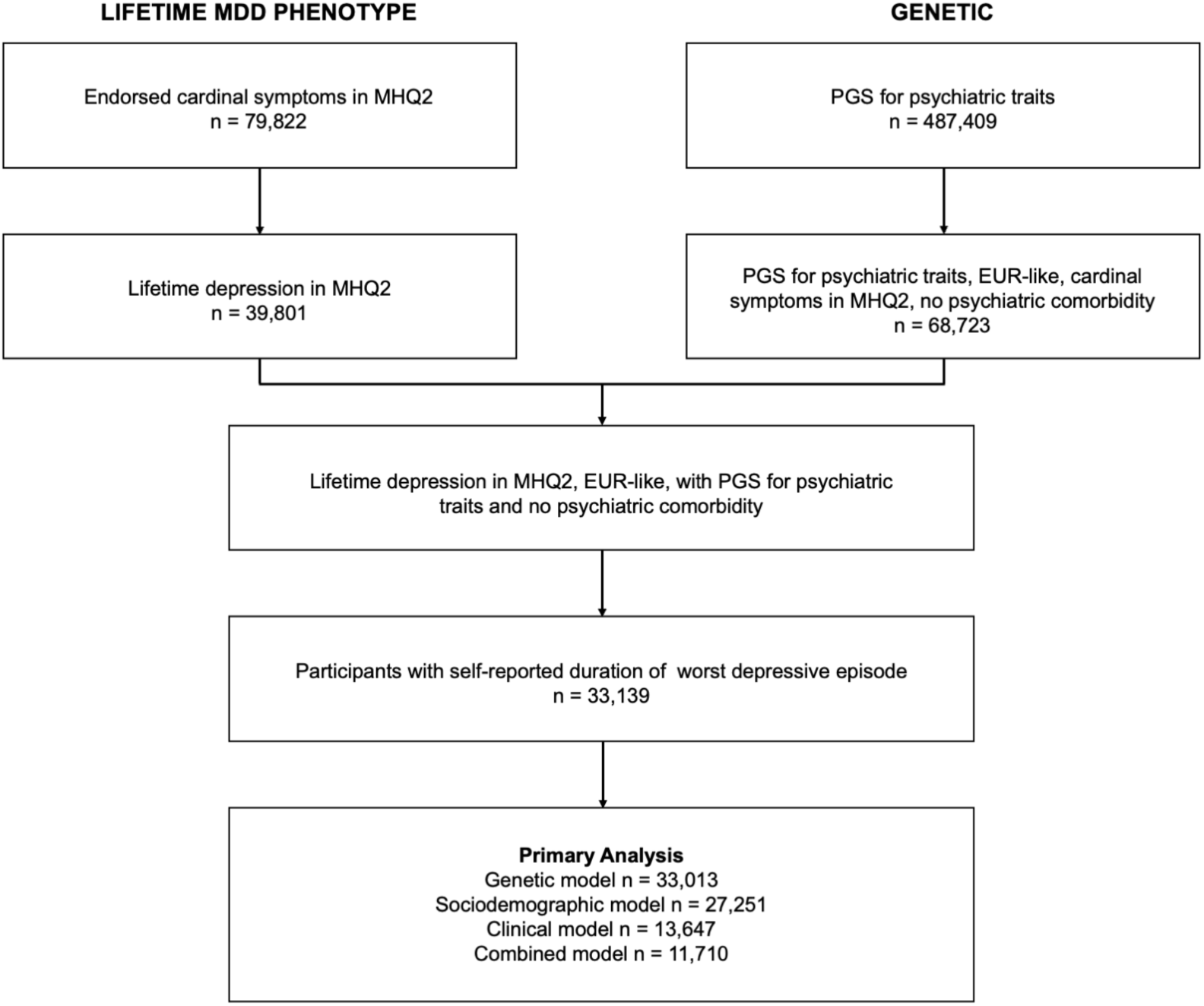
Flowchart illustrating sample derivation for primary analyses in UK Biobank participants. MDD: Major depressive disorder; MHQ2: Mental Health Questionnaire 2; PGS: polygenic scores.

Ethical approval for UKB was granted by the North-West Multi-centre Research Ethics Committee (approval number 11/NW/0382). All participation is voluntary, with the right to withdraw at any point, and written informed consent was obtained from all participants at baseline.

### Episode duration

The primary outcome was self-reported duration of worst depressive episode, where participants selected from six options: <1 month, 1-3 months, 3-6 months, 6-12 months, 12-24 months, or >24 months. Two binary outcomes were defined: 1) short episode duration (yes: <1 and 1-3 months combined; no: all other categories), and 2) long episode duration (yes: >24 months; no: all other categories). Distributions are reported in SI Section 2.

To contextualise the distributions of self-reported episode duration in UKB, we compared them with published estimates from Japanese study GLADS (Furukawa et al., 2000), and Dutch study NEMESIS (Spijker et al., 2002). Details are provided in SI Section 3.

### Risk factors

#### Clinical factors

Clinical variables relating to the participant’s depression history were derived from CIDI-SF responses. Lifetime number of depressive episodes was coded as a categorical variable, with participants reporting one (reference), 2-3, 4-5, 6 or more episodes, or selecting “too many to count/one episode runs into the next”. Further predictors included age of onset and years since the last episode, the latter calculated from reported age at last episode and the participant’s age at MHQ2 completion. A stressful trigger variable was coded as positive if the participant reported that their worst episode was precipitated by bereavement or a traumatic or stressful life event. Childhood trauma was assessed using the Childhood Trauma Screener (Grabe et al., 2012), a five-item measure capturing abuse and neglect. Items were dichotomised and summed to produce a total score ranging from 0 to 5, as described previously (Mutz et al., 2026). Family history of depression (yes/no) was based on participant report of depression among first-degree blood relatives. A depression subtype was derived; participants reporting both weight gain and hypersomnia during their worst episode were classified as increased weight-sleep; those reporting weight loss and insomnia or early waking were classified as decreased weight-sleep, others as neither. SI Section 1 provides UKB field codes and further details.

#### Sociodemographic factors

Sociodemographic variables included sex, body mass index (BMI), educational attainment, Townsend Deprivation Index (TDI), and neuroticism. Educational attainment was treated as a five-level categorical variable (university degree (reference), vocational, further, secondary, and none of the above). Neuroticism was measured using a derived summary score representing the total number of endorsed items across the 12 items of the neuroticism subscale of the Eysenck Personality Questionnaire-Revised Short Form (Eysenck et al., 1985), collected at baseline.

#### Genetic factors

Genome-wide genotyping is available for most UKB participants. Samples underwent standard quality control (QC) and imputation (Bycroft et al., 2018). PGS were calculated for six psychiatric conditions (depression, attention-deficit/hyperactivity disorder (ADHD), anorexia nervosa, bipolar disorder, obsessive compulsive disorder (OCD), and schizophrenia) and two selective serotonin reuptake inhibitor (SSRI) treatment response phenotypes of non-remission (SSRI_non-rem_) and percentage improvement (SSRI_perc_) (Hu et al., 2026). In brief, PGS were calculated using SBayesRC, and implemented within the GenoPred Pipeline (Pain et al., 2024; Zheng et al., 2024). Association testing was restricted to individuals genetically similar to European ancestry (EUR-like). Full details of GWAS summary statistics sources, QC procedures, and PGS computation are provided in SI Section 4.

### Analysis

#### Associations with episode duration

We tested associations between all variables and (a) short episode duration (0-3 months vs all other durations) and (b) long episode duration (>24 months vs all others) using binary logistic regression. All analyses were conducted using complete case analysis; participants with missing data or who reported “do not know” or “prefer not to answer” were excluded.

Three sets of analyses were conducted. First, 19 univariable models were fitted, one per variable. Second, three multivariable domain models were fitted: clinical, sociodemographic, and genetic, each containing all predictors from that domain simultaneously. Third, a combined model including all predictors simultaneously was fitted. All models included age at MHQ2 completion as a covariate. Sex was included as a covariate in univariable and in the clinical and genetic domain models, and as a predictor in the sociodemographic and combined models. Univariable PGS models and the genetic domain model were additionally adjusted for the first six genetic PCs and genotyping batch; in the combined model, each PGS was residualised on these covariates before entry alongside other predictors. Assessment centre was not included as a covariate (SI Section 5).

For the univariable and domain models for each outcome, Bonferroni-corrected p-values were obtained by multiplying observed p-values by the number of tests conducted (19, 8, 5, or 7 respectively). No multiple testing correction was applied to the combined model, as all predictors were entered simultaneously within a single model. To assess multicollinearity among predictors, variance inflation factors were calculated (SI Section 6).

Analysis code was written in Python 3 with assistance from Claude (Anthropic, https://claude.ai), between November 2025 and August 2026. Code was run using JupyterLab on the UKB Research Analysis Platform.

#### Exploratory prediction in a held-out sample

To explore out-of-sample prediction, we applied MHQ2-trained models to a held-out MHQ1 subset. Applying identical CIDI-SF phenotyping criteria to MHQ1 data, we identified participants who met the diagnostic inclusion criteria and excluded those who had contributed to the MHQ2 training model (n = 7,024; n = 3,646 with complete data on all predictors). Discriminative performance was evaluated using the area under the receiver operating characteristic curve (AUC). Training and test samples were compared on all model variables (SI Section 7).

#### Genetic analyses for episode duration

##### GWAS

GWAS of long and short MDD episodes were conducted in EUR-like ancestry participants (n = 40,858) across MHQ1 and MHQ2, as other ancestry groups were too small for well-powered genetic analyses. GWAS was performed using REGENIE v4.1 (Mbatchou et al., 2021), which accounts for relatedness, with covariates of genetic sex, age, assessment centre, genotyping platform, and the first six within-ancestry genetic PCs. Full details of sample QC, genotype QC, and GWAS parameters are provided in SI Section 8.

##### Heritability

Common-variant (SNP) heritability of each phenotype was estimated in unrelated EUR-like participants (n = 39,937) using GCTA’s GREML method (Yang et al., 2011), converted to the liability scale using UKB sample prevalence as an estimate of population prevalence (36% short, 13% long episodes; details in SI Section 8).

#### Associations with treatment outcomes

To examine whether episode duration was associated with treatment-seeking behaviour and perceived effectiveness, six binary treatment outcomes were derived from MHQ2 responses within the diagnosed sample. Questions covered help-seeking, prescribed medication, and therapeutic activities and whether they felt helpful, and the use of drugs or alcohol (SI Section 1). Logistic regression models for short and long episode duration were fitted, including covariates of age at MHQ2 completion and sex, with Bonferroni correction for 6 tests.

## Results

### Sample characteristics

Table 1 shows characteristics of participants completing MHQ2 by duration of worst episode.

**Table 1.** Characteristics of UK Biobank participants completing MHQ2.

| Episode duration |  | All | Short<br>0-3 months | Intermediate<br>3-24 months | Long<br>>24 months |
| --- | --- | --- | --- | --- | --- |
|  |  | N = 33,139<br>100% | N=11,853<br>35.77% | N = 16,955<br>51.16% | N = 4,331<br>13.07% |
| Age at MHQ2 completion | Mean (SD) | 67.65 (7.43) | 67.69 (7.45) | 67.64 (7.42) | 67.58 (7.43) |
| Sex | Female (%) | 69.6 | 65.1 | 73.0 | 69.0 |
|  | Male (%) | 30.4 | 34.9 | 27.0 | 31.0 |
| BMI | Mean (SD) | 27.15 (4.98) | 27.05 (4.76) | 27.08 (4.99) | 27.68 (5.50) |
| Education | University (%) | 45.1 | 47.4 | 45.1 | 38.8 |
|  | Vocational (%) | 11.5 | 11.6 | 11.1 | 12.6 |
|  | Further (%) | 14.9 | 14.5 | 15.1 | 15.0 |
|  | Secondary (%) | 23.3 | 21.9 | 23.5 | 26.4 |
|  | None of the above (%) | 5.2 | 4.6 | 5.1 | 7.2 |
| TDI | Mean (SD) | -1.54 (2.89) | -1.64 (2.84) | -1.55 (2.89) | -1.21 (3.03) |
| Neuroticism score | Mean (SD) | 5.64 (3.31) | 5.19 (3.22) | 5.63 (3.29) | 6.93 (3.32) |
| Number of episodes | 1 (%) | 33.5 | 41.2 | 32.9 | 14.5 |
|  | 2-3 (%) | 20.8 | 22.4 | 22.3 | 10.8 |
|  | 4-5 (%) | 11.8 | 12.9 | 12.6 | 5.9 |
|  | 6+ (%) | 6.9 | 7.5 | 7.1 | 4.6 |
|  | Too many/one runs into next (%) | 27.0 | 16.0 | 25.1 | 64.2 |
| Age of onset | Mean (SD) | 35.16 (16.09) | 35.88 (15.83) | 35.58 (16.00) | 31.42 (16.66) |
| Years since last episode | Mean (SD) | 12.56 (13.48) | 14.15 (14.30) | 12.44 (13.10) | 8.40 (11.60) |
| Stressful trigger | Yes (%) | 70.8 | 67.7 | 73.5 | 68.8 |
|  | No (%) | 29.2 | 32.3 | 26.5 | 31.2 |
| Childhood trauma score | Mean (SD) | 0.92 (1.22) | 0.82 (1.14) | 0.91 (1.21) | 1.28 (1.40) |
| Family history | Yes (%) | 44.5 | 40.6 | 45.7 | 50.1 |
|  | No (%) | 55.5 | 59.4 | 54.3 | 49.9 |
| Depression subtype | Inc. WS (%) | 6.2 | 4.9 | 6.3 | 9.5 |
|  | Dec. WS (%) | 39.7 | 38.5 | 42.3 | 33.0 |
|  | Neither (%) | 54.0 | 56.6 | 51.4 | 57.5 |

All: all participants, short: 0-3 months, intermediate 3-24 months (reference group), long: >24 months. BMI; Body mass index; MHQ2: Mental Health questionnaire 2; SD: standard deviation; TDI: Townsend deprivation index, higher values indicate more deprivation; WS: weight and sleep symptoms.

### Cross-study comparison of episode duration

The distribution of episode duration in UKB aligned with estimates from the Dutch epidemiological study (NEMESIS) and the Japanese clinical study (GLADS), suggesting that retrospective self-report of episode duration is a valid measure (Kaplan-Meier survival curve in SI Section 3). The proportion of participants whose episode lasted 6 months or less was 63.2% in UKB (MHQ2), 63.5% in NEMESIS and 43.5% in GLADS. At 24 months, the proportion remaining in an episode was 10.8% in UKB (MHQ2) and 16.7% in GLADS; NEMESIS showed a comparable proportion (20%) at its final 21-month data point.

### Episode duration is associated with clinical, sociodemographic, and genetic variables

A summary of the main results is presented below. Full results for all analyses are presented in SI Section 9, the domain-specific multivariable model results are presented in Figure 2, and Figure 3 summarises patterns of results across all three sets of analyses. Throughout this section, p-values reported for the univariable and domain-specific models are Bonferroni-adjusted (p_bon_), and p-values for the combined model are uncorrected.

**Figure 2.**
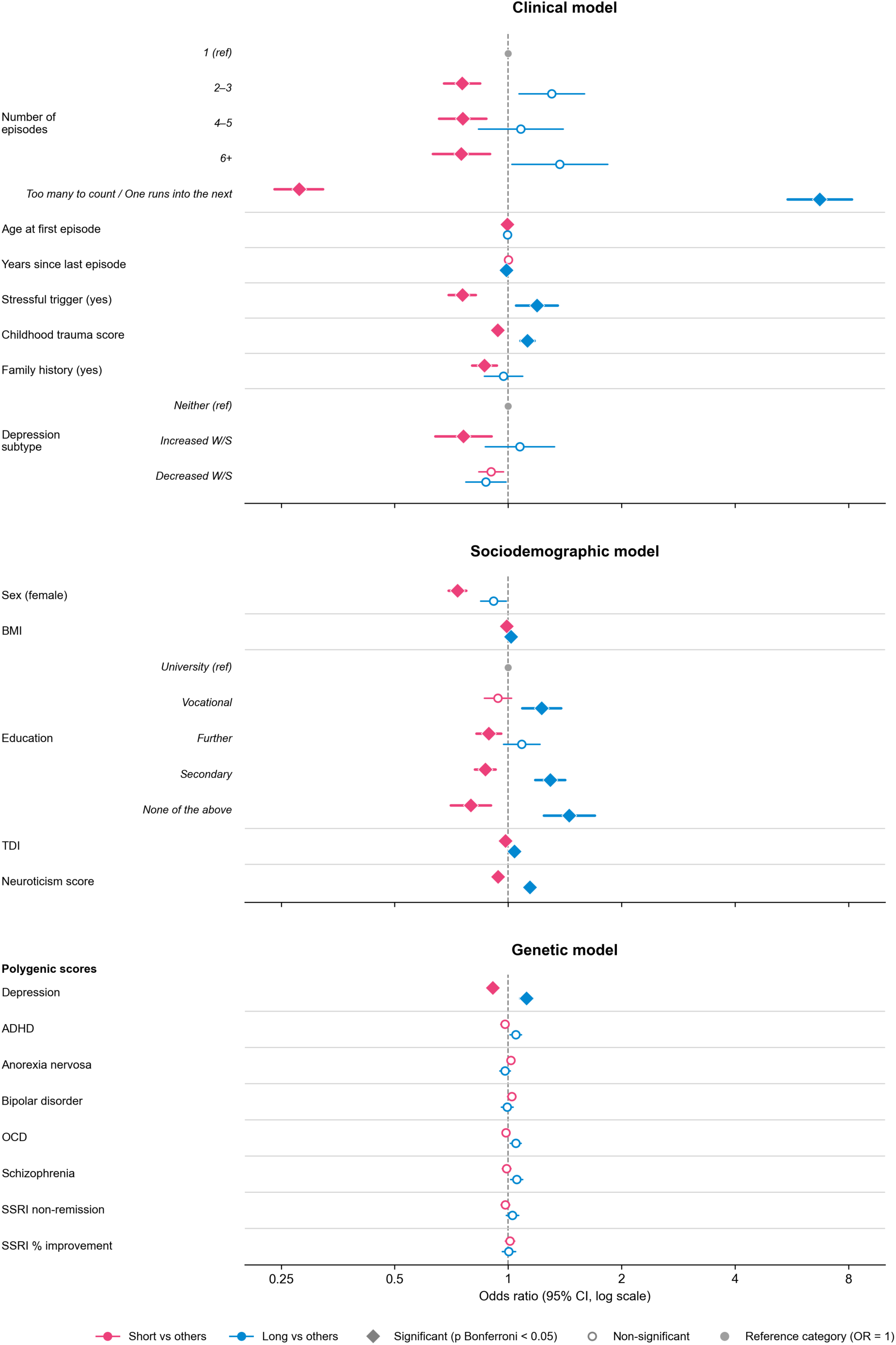
Associations between clinical factors, sociodemographic, and genetic predictors and duration of worst depressive episode in UK Biobank (domain-specific models). Odds ratios and 95% confidence intervals from logistic regression models comparing individuals with short (0 -3 months; pink) or long (>24 months; blue) worst depressive episode duration vs all others, for the clinical (top; N = 13,647), sociodemographic (middle; N = 27,251), and genetic (bottom; N = 33,013) models. Within each panel, all listed predictors were modelled simultaneously, and reference categories are marked. ADHD: attention-deficit/hyperactivity disorder; BMI: body mass index; OCD: obsessive compulsive disorder; TDI: Townsend Deprivation Index; WS: weight/sleep.

**Figure 3.**
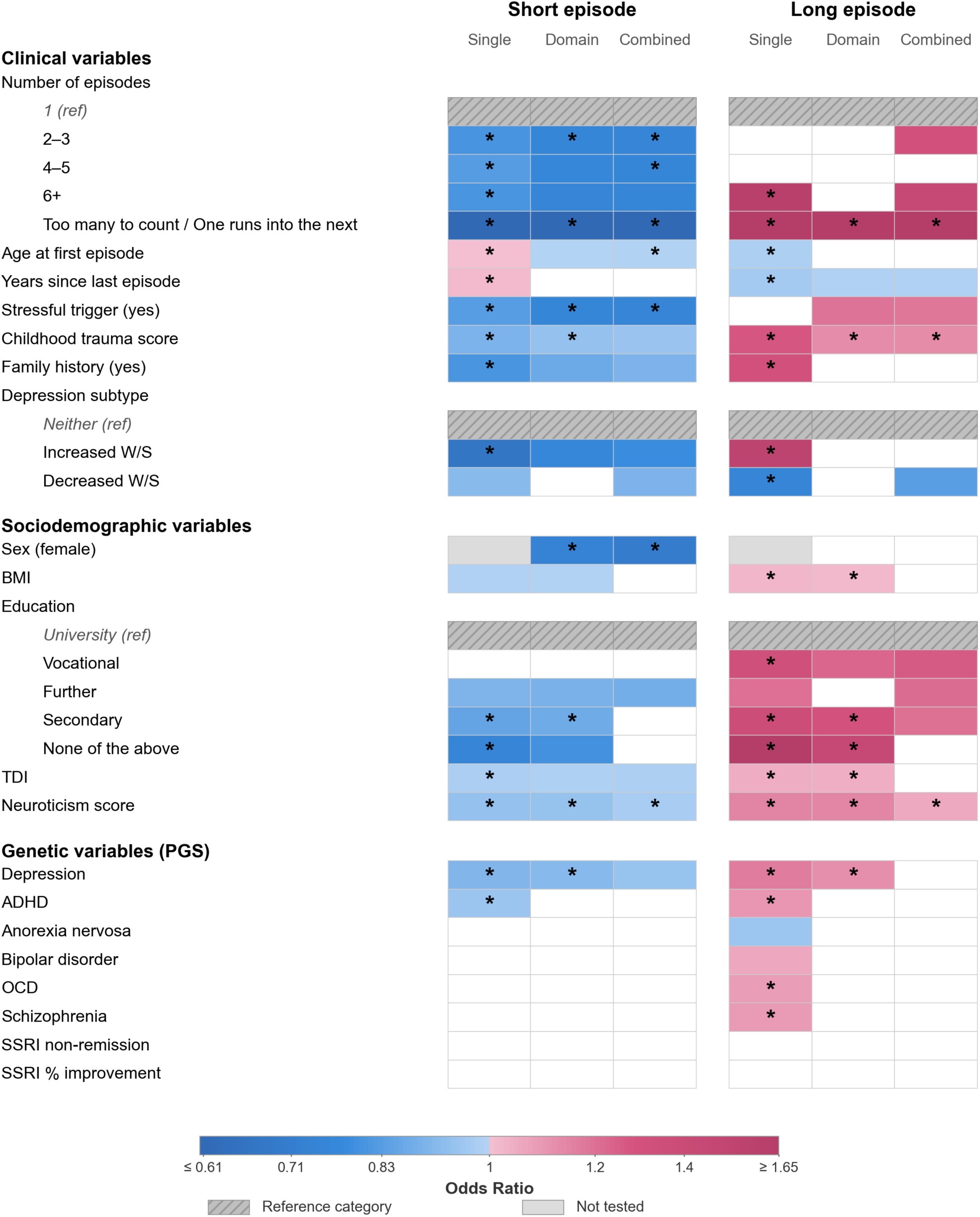
Associations between candidate predictors and short / long depressive episodes across univariable, domain-specific, and multivariable combined regression models. White cells are not significant; univariable and domain-specific models use Bonferroni-adjusted p-values, the combined model uses unadjusted p-values. Asterisks: pbon < 0.001. Solid grey cells indicate that sex was not included as a predictor in the univariable model. ADHD: attention-deficit/hyperactivity disorder; BMI: body mass index; OCD: obsessive compulsive disorder; PGS: polygenic score; SSRI: selective serotonin reuptake inhibitor; TDI: Townsend Deprivation Index.

#### Clinical variables

Clinical variables were strongly and consistently associated with episode duration across all models. In univariable analyses, having a higher number of lifetime episodes, higher childhood trauma scores, a family history of depression, and the increased weight/sleep subtype were each associated with reduced odds of a short episode (number of episodes: ORs = 0.339-0.828, all four contrasts; childhood trauma: OR = 0.893 [0.872-0.915]; family history: OR = 0.807 [0.771-0.846]; increased weight/sleep subtype: OR = 0.668 [0.596-0.749]; all p_bon_ < 0.001) and increased odds of a long episode (number of episodes: ORs = 1.609-7.564, two of four contrasts; childhood trauma: OR = 1.276 [1.239-1.315]; family history: OR = 1.310 [1.227-1.398]; increased weight/sleep subtype: OR = 1.543 [1.356-1.755]; all p_bon_ < 0.001). In the opposite direction, being older at onset and having a longer time since the last episode were associated with increased odds of a short episode (age of onset: OR = 1.003 [1.002-1.005]; years since last episode: OR = 1.014 [1.012-1.016]; both p_bon_ < 0.001) and reduced odds of a long episode (age of onset: OR = 0.982 [0.980-0.984]; years since last episode: OR = 0.967 [0.964-0.970]; both p_bon_ < 0.001). A stressful or traumatic trigger was associated with reduced odds of a short episode (OR = 0.830 [0.790-0.873], p_bon_ < 0.001) but not with long episodes; and the decreased weight/sleep subtype was associated with reduced odds of both short (OR = 0.917 [0.870-0.968], p_bon_ = 0.029) and long episodes (OR = 0.751 [0.695-0.811], p_bon_ < 0.001).

In the domain-specific multivariable model, more lifetime episodes (all four contrasts), childhood trauma, stressful trigger, family history, and the increased weight/sleep subtype remained associated with reduced odds of a short episode; more lifetime episodes (only the “too many to count/one episode runs into the next” contrast), and childhood trauma remained associated with increased odds of a long episode, and a longer time since the last episode remained associated with decreased odds. A stressful trigger became significantly associated with long episodes (OR = 1.193 [1.050-1.356], p_bon_ = 0.047).

In the combined multivariable model, more lifetime episodes, older age of onset, stressful trigger, childhood trauma, family history, and the increased weight/sleep subtype remained associated with reduced odds of a short episode. More lifetime episodes, stressful trigger, and childhood trauma remained associated with increased odds of long episodes while more recent episodes were associated with increased odds of long episodes.

#### Sociodemographic variables

Sociodemographic variables were also consistently associated with duration across models. In univariable models, higher BMI, lower educational attainment (versus university), greater deprivation, and a higher neuroticism score were each associated with reduced odds of a short episode (BMI: OR = 0.991 [0.987-0.996], p_bon_ < 0.003; education: ORs = 0.752-0.897, three of four contrasts; TDI: OR = 0.980 [0.972-0.988], p_bon_ < 0.001; neuroticism: OR = 0.939 [0.932-0.946], p_bon_ < 0.001) and increased odds of a long episode (BMI: OR = 1.024 [1.017-1.030]; education: ORs = 1.201-1.793, all four contrasts; TDI: OR = 1.045 [1.034-1.056]; neuroticism: OR = 1.148 [1.135-1.161]; all but one education contrast p_bon_ < 0.001).

In the domain-specific multivariable model, the same directional pattern held with attenuated effect sizes: higher BMI, lower educational attainment, greater deprivation, and higher neuroticism remained associated with reduced odds of a short episode and increased odds of a long episode, with one of the four education contrasts dropping below significance for long episodes. Female sex was associated with reduced odds of a short episode (OR = 0.734 [0.695-0.774], p_bon_ < 0.001), but not with long episodes.

In the combined model, associations attenuated further, and several predictors dropped below significance. For short episodes, deprivation, neuroticism, and female sex remained associated with lower odds, with only the further-education contrast surviving among education contrasts. For long episodes, neuroticism and educational attainment (three of four contrasts) remained associated with greater odds.

#### Genetic variables

In univariable models, higher depression PGS and ADHD PGS were associated with lower odds of short episodes (OR = 0.904 [0.882-0.926], p_bon_ < 0.001; and OR = 0.949 [0.928-0.972], p_bon_ < 0.001, respectively), and higher odds of long episodes (OR = 1.165 [1.125-1.206], p_bon_ < 0.001; and OR = 1.100 [1.064-1.137], p_bon_ < 0.001, respectively). OCD PGS (OR = 1.084 [1.050-1.119], p_bon_ < 0.001), schizophrenia PGS (OR = 1.091 [1.054-1.129], p_bon_ < 0.001) and bipolar disorder PGS (OR = 1.058 [1.023-1.093], p_bon_ = 0.017) were associated with higher odds of long episodes, while anorexia nervosa PGS (OR = 0.952 [0.924-0.982], p_bon_ = 0.036) were associated with lower odds of long episodes.

In the domain-specific genetic model, only depression PGS remained significantly associated with both short and long episode duration. In the combined multivariable model, depression PGS remained associated with short episode duration, though the effect size was attenuated. No genetic predictors were associated with long episode duration in the combined model.

### Exploratory prediction of episode duration

The domain-specific and combined models fitted in the primary MHQ2 sample were applied to a held-out subset of participants not used for model fitting (n = 3,646). For short episode duration, predictive performance was limited, ranging from near-chance for the genetic (AUC 0.536) and sociodemographic (AUC 0.550) models to modest discrimination for the clinical (AUC 0.603) and combined (AUC 0.601) models. Performance was meaningfully higher for long episode duration, with the clinical (AUC 0.710) and combined (AUC 0.705) models performing similarly. AUC values were broadly consistent with expectations given the training sample results, though training and validation samples differed on several demographic and clinical variables (SI Section 7).

### Genetic studies of episode duration

To identify novel genetic predictors of episode duration and assess its genetic architecture, we performed a GWAS of short episode duration, and of long episode duration. No genome-wide significant findings were identified for either outcome.

The liability-scale SNP-heritability was estimated to be 0.09 (SE = 0.04; p = 0.008) for long episode duration and 0.04 (SE = 0.02; p = 0.04) for short episode duration, where p-values are from one-sided likelihood ratio tests as implemented in GCTA. Common genetic variants thus explain a small but nominally significant proportion of variance in MDD episode duration. The point estimate was numerically higher for long than short episode duration, but a Wald test of the difference was not significant (p = 0.26); this comparison assumes independent estimates and does not account for sampling covariance between the two *h^2^* estimates, which were derived from the same sample. Manhattan and heritability plots are shown in SI Section 8.

### Association of episode duration with treatment outcomes

In the logistic regression, short episode duration was associated with significantly lower odds of all four treatment engagement outcomes: informing a professional, taking prescribed medication, using drugs, alcohol, or unprescribed medication, and undertaking therapeutic activities, but higher odds of reporting that therapeutic activities or SSRIs were helpful; this was reversed for long episode duration (Table 2).

**Table 2.**
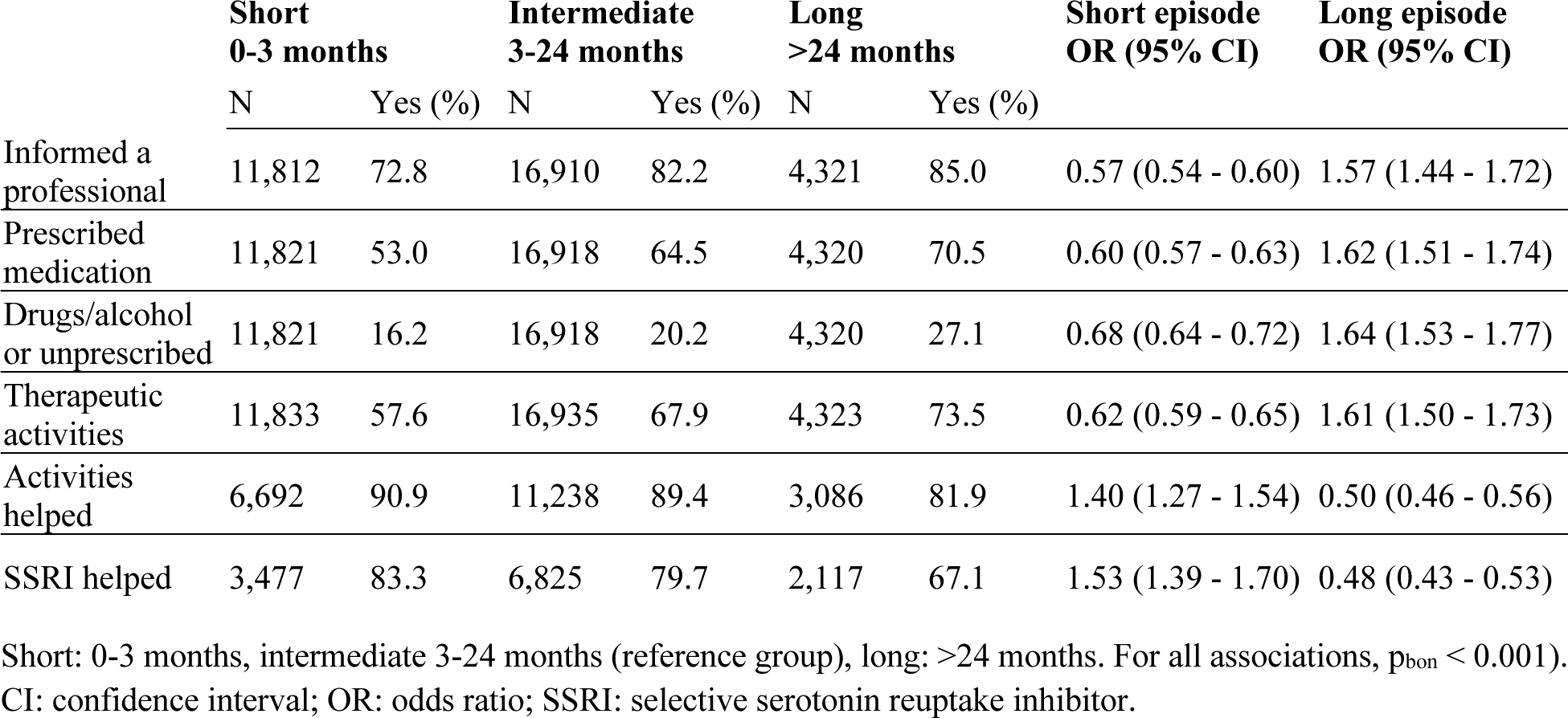
Treatment engagement and response distributions across short, intermediate, and long episode groups.

## Discussion

The duration of a depressive episode is an important component of the burden that depression places on individuals, healthcare services, and the economy. We present one of the first comprehensive analyses of episode duration, using self-reported data from UKB and focusing on two clinically meaningful endpoints: short episodes (under three months) and long episodes (over 24 months).

### Stress-related predictors of episode duration

Episode duration was robustly associated with childhood trauma, a stressful trigger, and a higher neuroticism score, each reducing the odds of a short episode and increasing the odds of a long one. Childhood maltreatment has been meta-analytically associated with recurrence, persistence, and poorer treatment response (Nanni et al., 2012; Xiong et al., 2026), and higher neuroticism with greater symptom severity and more persistent symptoms over time (Brown & Rosellini, 2011; van Eeden et al., 2019). Here, we show they are associated with episode duration when modelled alongside sociodemographic and genetic predictors.

One possible common thread is stress reactivity. Neuroticism is characterised by heightened reactivity to stress (Barlow et al., 2014); early-life adversity has been linked to lasting dysregulation of the hypothalamic-pituitary-adrenal axis (Bunea et al., 2017); and a stressful trigger acts on these systems. As this shared vulnerability may impair remission, measures of stress reactivity could be examined in future mechanistic work.

### Clinical history carries the signal

Clinical predictors were the most consistent across models, suggesting clinical features capture much of what is predictable about episode duration. Number of episodes was robustly associated with reduced odds of short and increased odds of long episodes. Recurrence and chronicity tend to co-occur, and a greater number of past episodes has been associated with greater severity, a more persistent course, and poorer treatment response (Kautzky et al., 2019). The strongest signal came from the “too many to count/one episode runs into the next” category, which may capture a chronic, unremitting course rather than a discrete episode count; associations were nonetheless graded across the 2-3, 4-5 and 6+ categories, supporting a dose-response relationship between recurrence and duration. As expected, a similar pattern of association was seen with an earlier age of onset and a shorter time since the last episode. Episode duration thus seems shaped by the broader trajectory of illness, consistent with evidence that recurrence, early onset, and recency load on a single dimension of chronicity (Pettit et al., 2009).

The weight/sleep subtypes broadly map onto atypical (increased weight/sleep) and melancholic (decreased) presentations of depression. The atypical-like subtype was associated with reduced odds of short episodes and increased odds of long episodes, consistent with a more chronic course. The melancholic-like subtype lowered the odds of both, suggesting that although melancholia is often considered more severe, these episodes cluster at intermediate durations.

Genetic and sociodemographic factors were also associated with duration. Higher BMI, lower educational attainment, and greater deprivation were associated with higher odds for long episodes, in line with studies linking socioeconomic disadvantage to a more persistent course of depression (Melchior et al., 2013). Depression PGS was the only consistent genetic signal, suggesting that higher genetic liability to depression is positively associated with episode duration. However, the association between depression PGS and long episodes attenuated in the combined model, suggesting it was partly mediated by clinical features that share genetic liability with depression, such as childhood trauma (Coleman et al., 2020). Common variants explained a small but nominally significant proportion of variance in both outcomes, indicating that episode duration is influenced by common genetic variation. No genome-wide significant loci were identified, as expected given the sample size and modest heritability. Sociodemographic and genetic information is available before a clinical history has accumulated, but discrimination in these domains was close to chance here, and considerably larger samples would be needed before either could contribute usefully to prediction.

### Long vs short episodes: A continuum of chronicity or a distinct subgroup?

The 24-month threshold used here aligns with the DSM-5 duration criterion for PDD (Schramm et al., 2020), though our long-episode group is not equivalent to a PDD diagnosis, as it was defined by self-reported worst episode duration without assessment of inter-episode recovery. Nonetheless, its key predictors (childhood trauma, recurrence, stressful trigger, and in univariable models, onset and family history) overlap with risk factors established for chronic depression (Hölzel et al., 2011; Nanni et al., 2012). This questions whether long episodes mark a discrete chronic subgroup or the extreme of a continuous dimension of chronicity. We show that most predictors acted in opposite directions for short and long episodes, as expected if both index a single underlying dimension. This mirroring is unlikely to be a result of how outcomes were defined; the melancholic-like subtype lowered odds of both short and long episodes, showing that non-opposing patterns were detectable. Depression PGS followed the same pattern and, being measured independently of symptom recall, indicates that common-variant liability is distributed across the duration continuum rather than concentrated in a discrete chronic subgroup. Our results therefore favour the view that long episodes represent the extreme of a continuum (Klein, 2008; Schramm et al., 2020). Sensitivity analyses to adjudicate between continuum and categorical constructs are priorities for future work.

### Clinical implications

Long episodes were considerably more predictable than short ones, and the features driving prediction, such as recurrence, childhood trauma, and stressful trigger, are ascertainable at presentation. Individuals with long episodes reported greater treatment engagement but were less likely to report benefit from medication or therapy, a pattern consistent with treatment resistance. The long-episode group is therefore a priority both for early identification and for developing more effective interventions, before persistent illness becomes entrenched.

### Limitations and future directions

UKB participants are on average healthier, more educated, and less deprived than the general UK population (Fry et al., 2017), and our analyses were restricted to European-like ancestry; replication in more diverse cohorts and multi-ancestry GWAS would improve both generalisability and power. The cross-sectional design means reverse causation cannot be excluded; a higher BMI may lengthen an episode, but a longer and more severe course may equally drive weight gain, and greater treatment engagement among those with long episodes partly reflects longer opportunity to seek help. Longitudinal data would be needed to establish direction. Retrospectively self-reported duration may be subject to recall bias (Horwitz et al., 2023); though reassuringly, such reports show reasonable validity against prospectively collected symptom data (Birk et al., 2020), and the UKB duration distribution corresponds closely to those from NEMESIS and GLADS.

## Conclusion

This study provides one of the first integrated analyses of clinical, sociodemographic, and genetic predictors of depressive episode duration, an understudied dimension of depression heterogeneity. It establishes that clinical history accounts for most of what can be predicted, and that long episodes were predicted more accurately than short ones. Patients at risk of a prolonged episode could therefore be identified early, and as this group also reported less benefit from treatment despite higher treatment-seeking, more intensive or different treatment could be considered from the outset.

## Acknowledgements

We gratefully acknowledge the UKB participants. This research was conducted under UKB project 82087. Claude (Anthropic, https://claude.ai), was used between November 2025 and August 2026 to draft and debug analysis code (see Methods) and to edit manuscript text for clarity; all AI-assisted content was reviewed by the authors, who take full responsibility for the final text.

## Ethical standards

The authors assert that all procedures contributing to this work comply with the ethical standards of the relevant national and institutional committees on human experimentation and with the Helsinki Declaration of 1975, as revised in 2008.

## Data availability

UK Biobank data are available to approved researchers through the UK Biobank Access Management System. Analysis code will be posted on Github (https://github.com/louisesophieschindler/Predictors-of-episode-duration) when the UK Biobank Research Analysis Platform reopens (estimated September 2026).

