## Supplementary Information for "Clinical, sociodemographic, and genetic predictors of depressive episode duration in the UK Biobank"

Louise S. Schindler[1, 2], Madhurbain Singh[1], Emerie Sheridan[1], Chris Wai Hang Lo[1], Michelle Kamp[1], Cathryn M. Lewis[1, 2]

[1] Social, Genetic, and Developmental Psychiatry Centre, King's College London, London, England, UK

[2] NIHR Biomedical Research Centre: Maudsley, South London and Maudsley NHS Foundation Trust, London, UK

**Section 1: UK Biobank field codes and descriptions**

**Table S1.** UK Biobank (UKB) field identifiers for all variables used in the primary (MHQ2) and validation (MHQ1) analyses from the UKB Data Showcase.

| Variable | MHQ2 | MHQ1 | Coding / Derivation |
|---|---|---|---|
| ***Outcome*** | | | |
| Duration of worst episode | 29030 | 20438 | Ordinal: 0-1, 1-3, 3-6, 6-12, 12-24, >24 months. |
| ***DSM-5 symptoms (used for diagnostic inclusion criteria)*** | | | |
| Prolonged sadness / depression | 29011 | 20446 | Cardinal symptom; binary (yes/no). |
| Prolonged loss of interest | 29012 | 20441 | Cardinal symptom; binary (yes/no). |
| Weight change (worst episode) | 29021 | 20536 | Binary (any change vs. no change). |
| Sleep change (worst episode) | 29022 | 20532 | Binary (yes/no). |
| Tiredness (worst episode) | 29018 | 20449 | Binary (yes/no). |
| Worthlessness (worst episode) | 29027 | 20450 | Binary (yes/no). |
| Concentration difficulty (worst episode) | 29026 | 20435 | Binary (yes/no). |
| Thoughts of death (worst episode) | 29029 | 20437 | Binary (yes/no). |
| Impact on normal roles (worst episode) | 29031 | 20440 | Binary (a lot / somewhat vs. not at all / not much). |
| ***Exclusion criteria variables*** | | | |
| Bipolar/depression diagnosis | 20126-0.0 | 20126-0.0 | Excluded if Bipolar I or II. |
| MHC diagnosis screen | 29000 | 29000 | Excluded if mania/hypomania, schizophrenia, or other psychosis. |
| ***Sociodemographic predictors*** | | | |
| Age at MHQ completion | Derived | Derived | Calculated from birth year (34), birth month (52), and date of MHQ completion (29183 / 20400). |
| Sex | 31 | 31 | Binary (female/male). Reference = male. |
| Body mass index (BMI) | 21001 | 21001 | Continuous; obtained from instance 0. |
| Educational attainment | 6138 | 6138 | Coded into categorical: none of the above, secondary (CSEs, GCSEs, or O-levels), further (A-levels or equivalent), vocational (NVQ/HND/HNC or professional qualifications), university (college or university degree). Reference = university. |
| Townsend Deprivation Index | 22189 | 22189 | Continuous; assigned at recruitment. |
| Neuroticism score | 20127 | 20127 | Continuous (0-12); UKB-derived summary score, obtained from instance 0. |

| | | | |
|---|---|---|---|
| ***Clinical predictors*** | | | |
| Lifetime number of episodes | 29033 | 20442 | Coded as categorical: 1, 2-3, 4-5, 6+, too many to count/one runs into the next. Reference = 1. |
| Age of onset | 29034 | 20433 | Continuous (years). |
| Years since last episode | Derived | Derived | Continuous; age at MHQ completion minus age at last episode (29036 / 20434). |
| Stressful trigger | 29013 | 20447 | Binary (yes/no). Worst episode precipitated by bereavement or stressful life event. Reference = no. |
| Childhood trauma score | 29076–29080 | 20487–20491 | Score 0-5; sum of five ACE items: felt loved (29076 / 20489), felt hated (29078 / 20487), physical abuse (29077 / 20488), sexual abuse (29079 / 20490), access to medical care (29080 / 20491). Each item was scored 0-4 (never vs often true); protective items (felt loved, access to medical care) were coded as trauma if ≤ 2, and abuse items (felt hated, physical abuse, sexual abuse) were coded as trauma if ≥ 1. |
| Family history of depression | 29001 | 29001 | Binary (yes/no). Coded yes if participant reported depression among first-degree relatives. Reference = no. |
| Depression subtype | Derived | Derived | Derived from weight change (29021 / 20536) and sleep items: trouble falling asleep (29023 / 20533), waking too early (29024 / 20535), and sleeping too much (29025 / 20534). Participants reporting weight gain and hypersomnia were classified as increased weight-sleep; weight loss with insomnia or early waking as decreased weight-sleep; all others as neither. Reference = neither. |
| ***Treatment variables*** | | | |
| Informed a professional | 29037 | - | Binary (yes/no). |
| Prescribed medication | Derived | - | Binary. Derived from 29038 (substances taken for depression). Coded 1 if the response contained "Medication prescribed to you for at least 2 weeks", 0 otherwise. |
| Drugs, alcohol, or unprescribed medication | Derived | - | Binary. Derived from 29038 (substances taken for depression). Coded 1 if the response contained "Drugs or alcohol" or "Unprescribed medication", 0 otherwise. |
| Therapeutic activities | Derived | - | Binary. Derived from 29047 (Activities undertaken to treat depression). Coded 1 if the response contained "Talking therapies" or "Other therapeutic activities", 0 otherwise. |
| Activities have helped | 29048 | - | Binary. Coded 1 if "Yes, at least a little", 0 if "No". |
| Antidepressants have helped | Derived | - | Binary. Composite variable derived from four individual SSRI helpfulness items: citalopram (29040), fluoxetine (29041), sertraline (29042), paroxetine (29043). Derivation of the composite variable is described elsewhere (Kamp et al., 2025). |

*Derived* indicates that the variable was calculated from multiple fields (see Coding/Derivation column). ACE: adverse childhood experience; MHC: mental health conditions.

**Section 2: Distribution of long and short episodes**

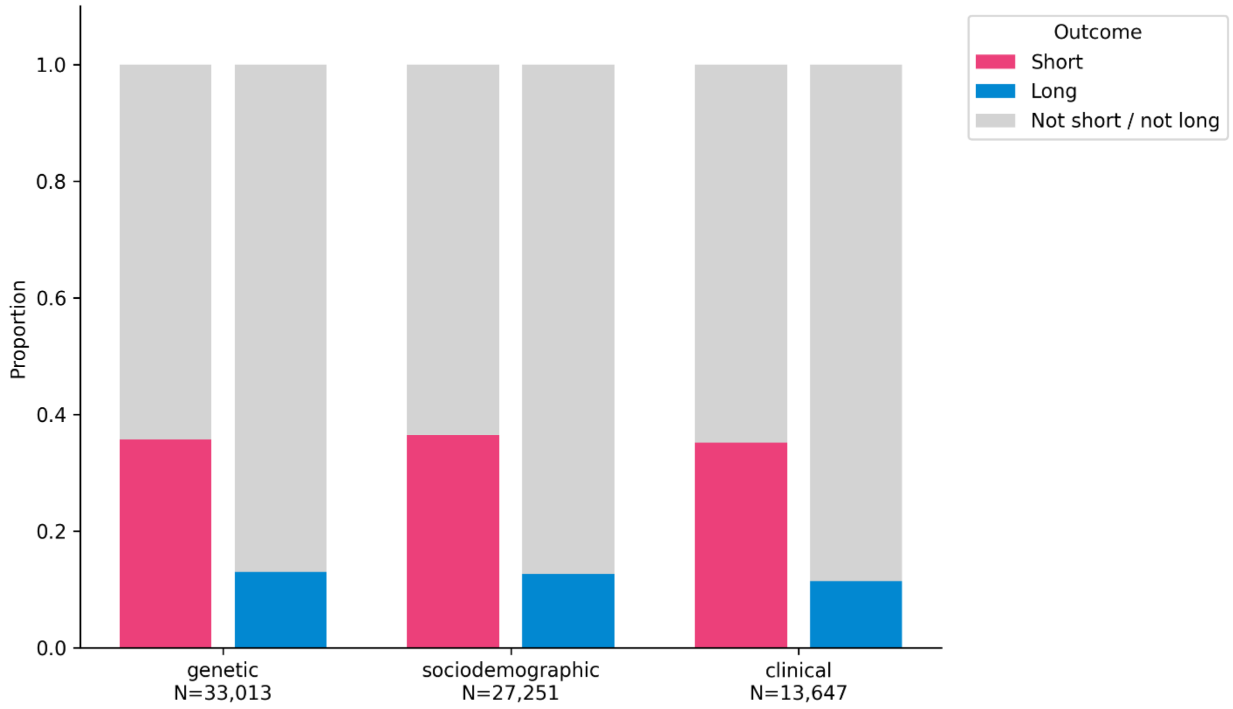


**Figure S1. Distribution of episode duration categories across model-specific analytic samples.** Participants were classified as having a short (0-3 months; pink), long (>24 months; blue), or not short/not long (grey) episode duration.

**Section 3: Cross-study comparison of episode duration**

We compared the distributions of self-reported episode duration in UKB with published estimates from the Group for Longitudinal Affective Disorders Study (GLADS), a clinical study from Japan (Furukawa et al., 2000), and the Netherlands Mental Health Survey and Incidence Study (NEMESIS) (Spijker et al., 2002). Episode duration data were digitised from published figures using https://automeris.io/, and Kaplan-Meier curves of episode duration were plotted alongside for comparison with UKB (Figure S2).

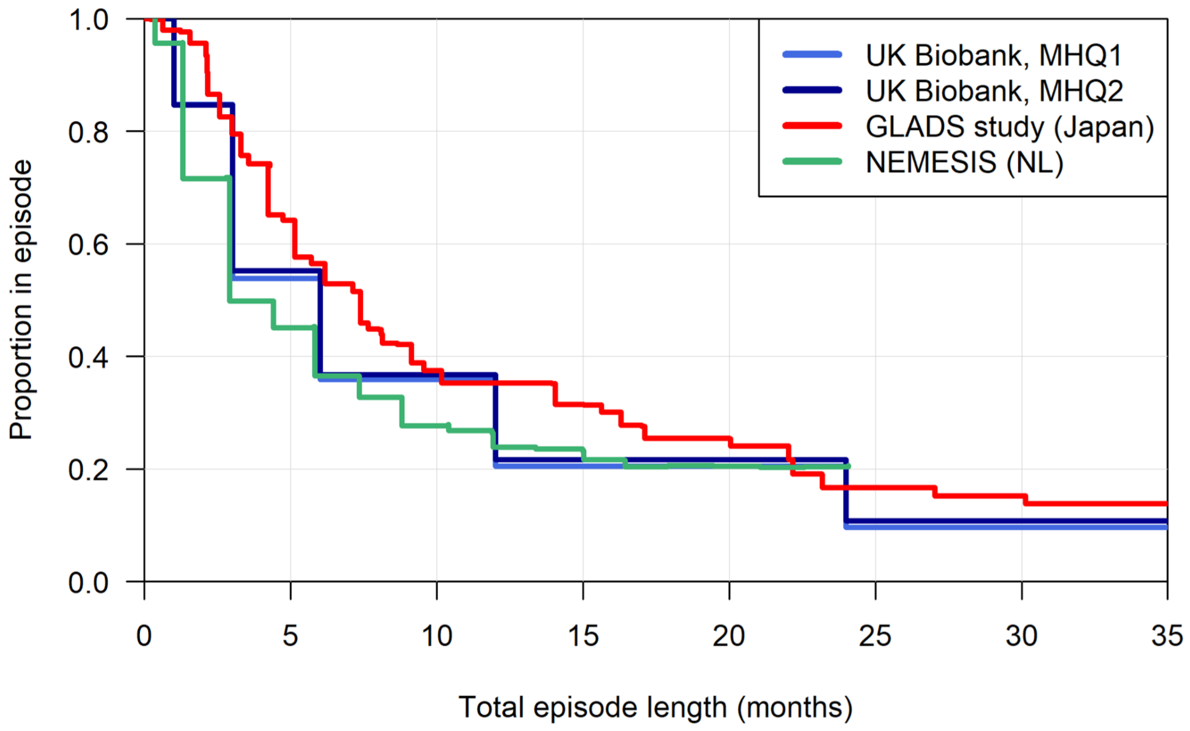


**Figure S2. Kaplan-Meier survival curves.** Plot shows the proportion of participants remaining in a depressive episode over time across four samples: UK Biobank MHQ1 and MHQ2, the GLADS study (Japan), and NEMESIS (Netherlands). MHQ: Mental Health Questionnaire.

### Section 4: Polygenic scores

Genome-wide genotyping is available for all UKB participants. Samples underwent standard quality control (QC) and imputation. Polygenic scores (PGS) were calculated for several psychiatric conditions: major depressive disorder (MDD), attention-deficit/hyperactivity disorder (ADHD), anorexia nervosa, bipolar disorder, obsessive compulsive disorder (OCD), schizophrenia, and two SSRI-treatment response phenotypes: non-remission ($SSRI_{non-rem}$) and percentage improvement ($SSRI_{perc}$), using GWAS summary statistics from the Psychiatric Genomics Consortium (Table S2). In brief, $SSRI_{perc}$ and $SSRI_{non-rem}$ were calculated as previously described (Pain et al., 2022); $SSRI_{perc}$ was calculated as 100 * (baseline depression score − final depression score) / baseline depression score, with higher values indicating greater improvement; remission was defined as symptom reduction to a pre-specified threshold for the relevant rating scale, with those failing to reach this threshold classified as non-remitters.

**Table S2.** Source of GWAS summary statistics.

| Phenotype | PMID | Authors | N | Case N | Control N |
|---|---|---|---|---|---|
| MDD | 39814019 (excl. UKB + 23andMe) | (Adams et al., 2025) | 1,639,572 | 357,636 | 1,281,936 |
| ADHD | 36702997 | (Demontis et al., 2023) | 225,534 | 38,691 | 186,843 |
| Anorexia nervosa | 31308545 | (Watson et al., 2019) | 68,684 | 16,224 | 52,460 |
| Bipolar disorder | 39843750 | (O'Connell et al., 2025) | 876,091 | 66,494 | 809,597 |
| OCD | 40360802 | (Strom et al., 2025) | 1,011,601 | 22,717 | 988,884 |
| Schizophrenia | 35396580 | (Trubetskoy et al., 2022) | 320,404 | 76,755 | 243,649 |
| SSRI non-remission | NA | (Hu et al., 2026) | 3,872 | 2,371 | 1,498 |
| SSRI % improvement | NA | (Hu et al., 2026) | 3,887 | 3,887 | 0 |

Summary statistics were used to develop six psychiatric disorder-related polygenic scores and two antidepressant response-associated trait scores. ADHD: attention-deficit/hyperactivity disorder; MDD: major depressive disorder; OCD: obsessive compulsive disorder; SSRI: selective serotonin reuptake inhibitor; UKB: UK Biobank.

PGS were calculated using SBayesRC (Zheng et al., 2024), implemented within the GenoPred Pipeline (Pain et al., 2024). In GenoPred, GWAS summary statistics were quality-controlled by filtering duplicated SNPs, SNPs with low imputation quality (INFO < 0.9) and SNPs with out-of-bound p-values ($p < 0$ or $p > 1$). SNPs with minor allele frequencies (MAF) < 0.01 were also filtered. Quality-controlled GWAS summary statistics were further matched to the 1000 Genomes Phase 3 and Human Genome Diversity Project (HGDP) genomic reference panels, consisting of 1,204,449 variants for 3,313 individuals [https://opain.github.io/GenoPred/index.html]. Strand-flipping of SNPs was performed by matching alleles in summary statistics to the reference panel. To ensure the robustness of PGS (the computation of which is based on imputed variants), only SNPs present in the well-imputed HapMap3 variant list were retained for downstream PGS computation. SNPs with MAF differences > 0.2 between the genomic reference panel and GWAS summary statistics were also removed to ensure consistency in allele frequencies between the genome reference panel and the population on which the original discovery GWAS was performed.

SBayesRC is a Bayesian-based polygenic scoring method that leverages functional annotations to better account for linkage disequilibrium (LD) of SNPs, and subsequently better estimate SNP effect sizes from GWAS summary statistics (Zheng et al., 2024). It has demonstrated superior predictive performance across complex traits when applied to large European GWAS summary statistics (Pain, 2025). In the GenoPred pipeline, PGS were computed by SBayesRC using SNP effect sizes from the quality-controlled GWAS summary statistics, matched to an LD reference from a random sample of 20,000 unrelated EUR participants in UKB (Zheng et al., 2024). The PGS were corrected for genetic principal components (PCs) projected from the reference panels (1KG and HGDP) on a continuous scale, to account for systematic population structure in PGS distributions.

These PGS were further used for association testing with episode length variables listed below, only on European individuals identified using principal components projected from the pan-UKB reference panel (Karczewski et al., 2025). Covariates included the first six within-ancestry genetic principal components (PCs), age at MHQ2 completion, sex, and genotyping microarray platform (either the UK Biobank Axiom array or the UKB BiLEVE Axiom array) (Bycroft et al., 2018).

**Section 5: Assessment centre associations**

To assess whether assessment centre required inclusion as a covariate, chi-square tests of independence were conducted between assessment centre (22 UKB sites) and each outcome variable. Assessment centre was not significantly associated with short episode status ($\chi^2$ (21) = 21.37, $p = 0.436$) or long episode status ($\chi^2$ (21) = 19.34, $p = 0.563$) and was therefore not included in any models.

### Section 6: Multicollinearity amongst variables

Variance inflation factors (VIF) were calculated for each predictor within the combined model to assess multicollinearity. Conventional thresholds treat VIF values above 5 as indicative of problematic multicollinearity and values above 10 as serious (Kim, 2019). All VIF values were below 2.3 (Table S3), well within acceptable limits. The estimates are therefore not meaningfully inflated by shared variance, supporting the interpretability of the individual predictor effects reported in the main analysis.

**Table S3.** Variance inflation factors (VIF) for all predictors in the combined model.

| **Predictor** | **VIF** | **Predictor** | **VIF** | **Predictor** | **VIF** |
|---|---|---|---|---|---|
| ***Genetic (PGS)*** | | ***Sociodemographic*** | | ***Clinical*** | |
| MDD | 1.28 | Sex | 1.01 | Number of episodes | |
| ADHD | 1.15 | BMI | 1.02 | 2-3 | 1.72 |
| Anorexia nervosa | 1.06 | Education | | 4-5 | 1.74 |
| Bipolar disorder | 1.22 | Vocational | 1.12 | 6+ | 1.56 |
| OCD | 1.08 | Secondary | 1.14 | Too many to count | 2.28 |
| Schizophrenia | 1.19 | Further | 1.17 | Age at onset | 1.93 |
| SSRI non-remission | 1.43 | None of the above | 1.10 | Years since last episode | 1.98 |
| SSRI % improvement | 1.43 | TDI | 1.02 | Stressful trigger | 1.07 |
| | | Neuroticism score | 1.02 | Childhood trauma score | 1.08 |
| | | | | Family history | 1.07 |
| | | | | Depression subtype | |
| | | | | Increased weight/sleep | 1.09 |
| | | | | Decreased weight/sleep | 1.07 |

VIF values are identical for the short and long episode duration outcomes as both models share the same predictor set. ADHD: attention-deficit/hyperactivity disorder; BMI: body mass index; MDD: major depressive disorder; OCD: obsessive compulsive disorder; PGS: polygenic scores; SSRI: selective serotonin reuptake inhibitor; TDI: Townsend Deprivation Index.

### Section 7: Comparison between MHQ2 training and held-out samples

Comparisons between the combined MHQ2 training sample (N = 11,710) and the held-out sample (N = 3,646) on all model variables are reported in Table S4. The training sample was older on average, had more female participants, had a higher BMI, higher educational attainment, and higher neuroticism scores. They had higher PGS for MDD and bipolar disorder, higher number of lifetime episodes, more years since the last episode, higher proportion of family history, and higher proportions in the decreased and increased weight/sleep subtypes. They had a lower proportion of stressful or traumatic triggers.

**Table S4.** Comparison of MHQ2 training and MHQ1 held-out samples.

| | **MHQ2 (N = 11,710)** | **MHQ1 (N = 3,646)** | **Statistic** | ***p*** |
|---|---|---|---|---|
| ***Genetic variables (PGS)*** | | | | |
| MDD | 0.05 (0.94) | -0.01 (0.96) | t = 3.03 | **0.002** |
| ADHD | 0.01 (0.97) | 0.00 (0.95) | t = 0.19 | 0.851 |
| Anorexia nervosa | 0.34 (1.05) | 0.36 (1.05) | t = -1.10 | 0.270 |
| Bipolar disorder | -0.05 (0.97) | -0.12 (0.98) | t = 3.93 | **<0.001** |
| OCD | 0.03 (1.01) | -0.01 (1.01) | t = 1.90 | 0.058 |
| Schizophrenia | -0.31 (0.94) | -0.30 (0.95) | t = -0.57 | 0.571 |
| SSRI non-remission | 0.13 (1.02) | 0.16 (1.03) | t = -1.57 | 0.115 |
| SSRI % improvement | 0.05 (0.97) | 0.03 (0.96) | t = 1.25 | 0.210 |
| ***Sociodemographic variables*** | | | | |
| Age at MHQ completion | 67.31 (7.34) | 62.83 (7.17) | t = 32.39 | **<0.001** |
| Sex, % female | 69.0 | 66.6 | $\chi^2$ = 7.30 | **0.007** |
| BMI | 27.04 (5.01) | 26.82 (4.58) | t = 2.41 | **0.016** |
| Education | | | $\chi^2$ = 17.55 | **0.002** |
| University | 48.7 | 46.5 | | |
| Vocational | 10.4 | 11.2 | | |
| Secondary | 22.0 | 22.2 | | |
| Further | 15.0 | 14.8 | | |
| None of the above | 3.9 | 5.3 | | |
| TDI | -1.71 (2.77) | -1.73 (2.79) | t = 0.43 | 0.667 |
| Neuroticism score | 5.31 (3.29) | 4.94 (3.13) | t = 5.94 | **<0.001** |
| ***Clinical variables*** | | | | |
| Number of episodes | | | $\chi^2$ = 103.45 | **<0.001** |
| 1 | 38.1 | 43.3 | | |
| 2-3 | 23.3 | 27.2 | | |
| 4-5 | 11.9 | 10.3 | | |
| 6+ | 6.6 | 4.5 | | |
| Too many to count | 20.1 | 14.8 | | |
| Age of onset | 36.51 (15.89) | 36.86 (14.52) | t = -1.19 | 0.232 |
| Years since last episode | 12.86 (13.20) | 12.12 (11.74) | t = 3.02 | **0.003** |

| | | | | |
|---|---|---|---|---|
| Stressful trigger, % yes | 72.8 | 75.9 | $\chi^2 = 13.32$ | **<0.001** |
| Childhood trauma score | 0.91 (1.23) | 0.88 (1.15) | t = 1.23 | 0.220 |
| Family history, % yes | 43.2 | 35.2 | $\chi^2 = 72.61$ | **<0.001** |
| Depression subtype | | | $\chi^2 = 26.06$ | **<0.001** |
| Increased weight/sleep | 5.8 | 4.6 | | |
| Decreased weight/sleep | 41.4 | 37.9 | | |
| Neither | 52.9 | 57.5 | | |

Means and standard deviations (SD) are reported for continuous variables. Percentage distributions are reported for categorical and binary variables. Group differences for continuous variables were tested using independent-samples t-tests; categorical variables were tested using chi-square tests. ADHD: attention-deficit/hyperactivity disorder; BMI: body mass index; MDD: major depressive disorder; MHQ: Mental Health Questionnaire; OCD: obsessive compulsive disorder; PGS: polygenic scores; SSRI: selective serotonin reuptake inhibitor; TDI: Townsend Deprivation Index.

Figure S3 shows the Receiver Operating Characteristic (ROC) curve for the domain-specific and combined models predicting episode duration in the held-out sample.

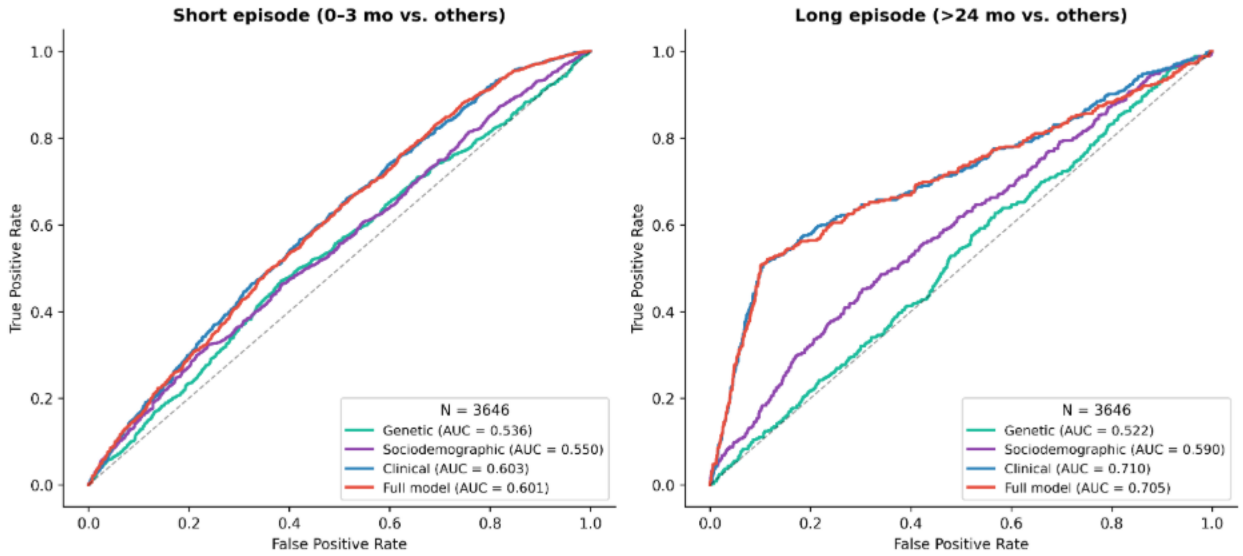


**Figure S3. Receiver Operating Characteristic (ROC) curves for domain-specific and combined models predicting episode duration in the held-out sample.** Left panel: short episode, right panel: long episode. N = 3,646. ROC curves for the genetic (teal), sociodemographic (purple), clinical (blue), and full combined (red) models, evaluated on participants phenotyped from MHQ1 responses and not used for model fitting.

**Section 8: GWAS and SNP-based heritability**

*Genetic Ancestry Groupings and Within-Ancestry Genetic Principle Component Analysis*

We used the genetic ancestry group assignments (Data-Field 30079) provided by the Pan-UKB Project (Karczewski et al., 2025). These ancestry assignments are based on the genetic similarity of UKB participants to six reference population groups in the joint 1000 Genomes Project Phase-3 and Human Genome Diversity Project (KGP-HGDP): Admixed American (AMR), African (AFR), Central/South Asian (CSA), East Asian (EAS), European (EUR), and Middle Eastern (MID) (Koenig et al., 2024). The assigned ancestry group distribution is shown in Table S5.

**Table S5.** Genetic Ancestry Group Distribution in the UK Biobank.

| Ancestry Group | Total N | Unrelated Subset |
|---|---|---|
| AFR-like | 6,740 | 6,145 |
| AMR-like | 992 | 946 |
| CSA-like | 9,056 | 8,215 |
| EAS-like | 2,778 | 2,624 |
| EUR-like | 426,454 | 353,418 |
| MID-like | 1,620 | 1,512 |
| NA (unassigned) | 54,296 | 33,794 |

We computed within-ancestry genetic principal components (PCs) using *FlashPCA2* (Abraham et al., 2017). We used Data-Field 22020 to identify the unrelated subset of participants (N = 406,654) previously used for genetic principal components analysis (PCA) in the UKB (Table S5). We then created unrelated ancestry-specific subsets of the genotyped microarray data and performed genetic variant and sample QC using *PLINK1.9* (Chang et al., 2015; Purcell et al., 2007). Within each ancestry subset, we retained SNPs with missingness rate < 2%, MAF >0.01, and Hardy-Weinberg equilibrium (HWE) p-value >1e-8. We excluded genomic regions with known long-range LD (Price et al., 2008) and pruned the remaining SNPs with LD-r2 >0.1 within a window of 1,500 kb (--indep-pairwise 1500 150 0.1). The ancestry-specific lists of QC'ed, semi-independent SNPs were used to create unrelated and related subsets of each ancestry group, along with additional QC to exclude samples with missingness >10%.

Using *FlashPCA2* we first computed the first 20 genetic PCs and the corresponding SNP weights in the unrelated subset of each ancestry group and then used the SNP weights to project the related samples onto the PCs. For quality checks, we also examined the root mean squared error (RMSE) for each set of PCA, which was estimated to be < 2e-8 in all ancestry groups.

Genetic analyses (GWAS and polygenic score analyses) were limited to EUR-like ancestry participants because of the small sample sizes of other ancestry groups.

*Genome-Wide Association Study (GWAS)*

GWAS of long and short MDD episodes were performed using *REGENIE* v4.1 to include related participants (Mbatchou et al., 2021).

Sample Filtering

For both phenotypes, GWAS was limited to EUR-like ancestry participants. We excluded participants with a potential sex chromosome aneuploidy (Data-Field 22019) or a mismatch between the genetically inferred sex (Data-Field 22001) and the self-reported sex (Data-Field 31). Further sample QC was done based on the genotype microarray data and whole-genome sequence (WGS) data, as described below.

Microarray Genotype Data QC

For *REGENIE* Step 1, we selected microarray genotyped, high-quality SNPs (Data-Field 22418) with MAF >0.01, minor allele count (MAC) >50, missingness rate < 2%, and HWE p-value >1e-15 within ancestry group, using *PLINK2* (Chang et al., 2015). Using the QC'ed SNP list, we excluded participants with sample genotyping call rate < 95%.

WGS Genotype Data QC

For *REGENIE* Step 2, we QC'ed the ML-corrected DRAGEN WGS genotype calls (Data-Field 24308) to retain variants with MAF > 0.01, missingness rate < 5%, and HWE p-value >1e-15 within ancestry group, using *PLINK2*. The WGS QC was done stratified by ancestry groups for all UKB participants, rather than limited to those with the phenotype of interest. All participants had samples with genotyping call rate > 95%, so no further participants were excluded based on sample genotyping missingness.

The QC'ed sample size with both microarray and WGS data was 40,858, comprising 5,257 cases and 35,601 controls for long episodes, and 14,739 cases and 26,119 controls for short episodes.

REGENIE

For each phenotype, we applied a logistic regression model in both steps of *REGENIE*, with covariates of sex (Data-Field 31), age at the baseline assessment (Data-Field 21003_i0), assessment centre (Data-Field 54_i0), microarray platform (either Axiom or BiLEVE; derived from the genotyping batch in Data-Field 22000), and the first 6 within-ancestry genetic PCs.

*REGENIE* Step-1 implemented a whole-genome ridge logistic regression model using the QC'ed autosomal microarray genotype data, yielding a genetically predicted case probability for each participant. Here, we used a block size (--bsize) of 1000.

In Step 2 GWAS, a SNP-wise logistic regression was conducted using the QC'ed WGS genotypes, conditional on the genetic prediction from Step 1. Here, we applied *REGENIE*'s approximate Firth correction (Mbatchou et al., 2021) to p-values <0.01 to minimise false positives under imbalanced case-control ratios, along with a back-correction to compute the Firth-corrected standard errors. *REGENIE* applies a leave-one-chromosome-out (LOCO) algorithm in Step 2, such that the SNP-trait associations on a given chromosome are estimated conditional on the Step-1 prediction excluding that chromosome. We also analysed chromosome X genotypes in Step 2, wherein *REGENIE* examines males as diploids (i.e., with genotypes as either 0 or 2).

GWAS Results

No variant reached genome-wide significance ($p < 5 \times 10^{-8}$) for either phenotype. Manhattan plots for both GWAS are shown in Figure S4.

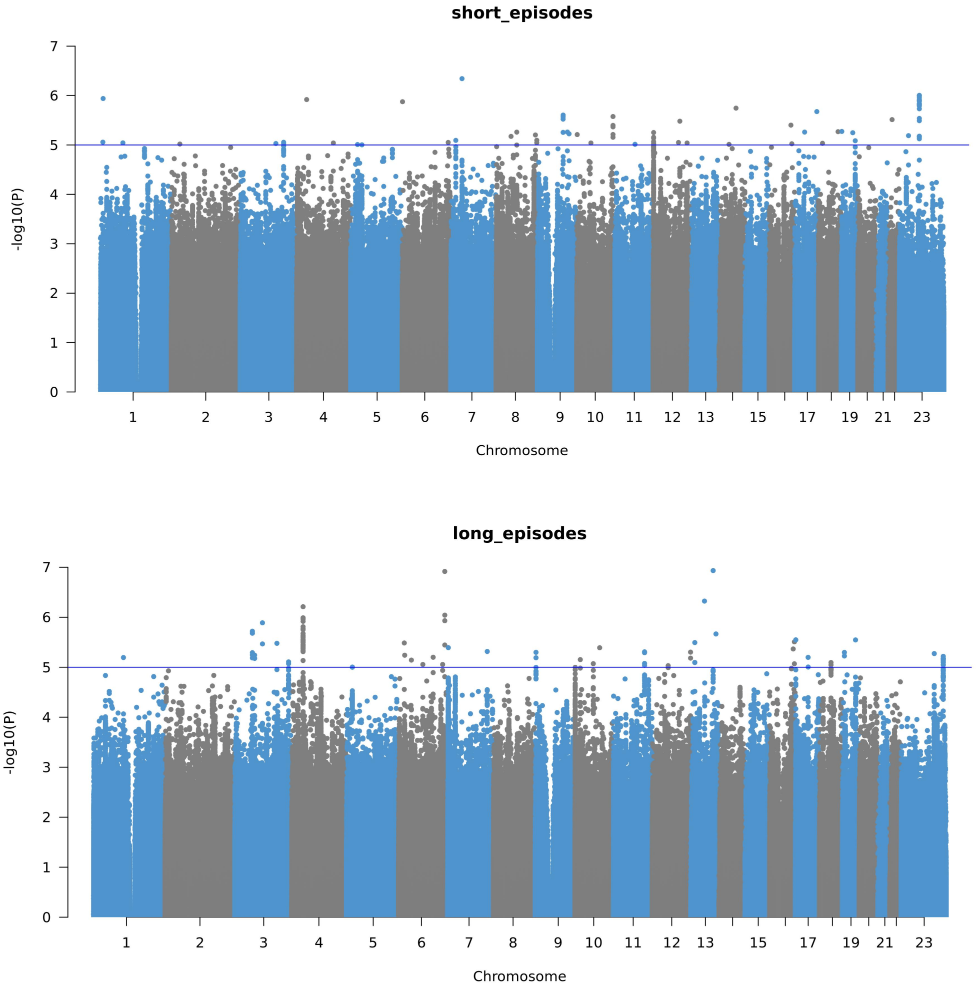


**Figure S4. Manhattan plots of the genome-wide association studies of episode duration in UK Biobank participants.** (A) Short episode duration (14,739 cases, 26,119 controls). (B) Long episode duration (5,257 cases, 35,601 controls).

*SNP Heritability*

Methods

We estimated the common-variant heritability (SNP heritability) of each phenotype in the EUR-like participants, using *GCTA* v1.95.1 (Yang et al., 2011). We used the QC'ed autosomal microarray genotypes, as in *REGENIE* Step 1, to construct a genetic relatedness matrix (GRM).

The GRM was then adjusted for incomplete LD between the genotyped SNPs and the causal variants, using the --grm-adj 0 flag. We excluded participants with estimated genetic relatedness greater than 0.05 in the GRM. The remaining unrelated subset (N = 39,937) comprised 5,141 cases and 34,796 controls for long episodes, and 14,378 cases and 25,559 controls for short episodes.

Observed-scale SNP heritability was then estimated in the unrelated participants using the single-component GREML (genomic restricted maximum likelihood) model with the same covariates as in the GWAS. The observed-scale heritability and standard error estimates were converted to the liability scale (Lee et al., 2012), assuming the population prevalence to be same as the sample prevalence (i.e., 13% and 36% for long and short episodes, respectively). The corresponding p-values were calculated within GCTA as $0.5 \times P(\chi^2_1 \geq$ Likelihood Ratio Test (LRT)).

Results

The liability-scale SNP-heritability was estimated to be 0.09 (SE = 0.04; p = 0.008) for long episode duration and 0.04 (SE = 0.02; p = 0.04) for short episode duration (Figure S5).

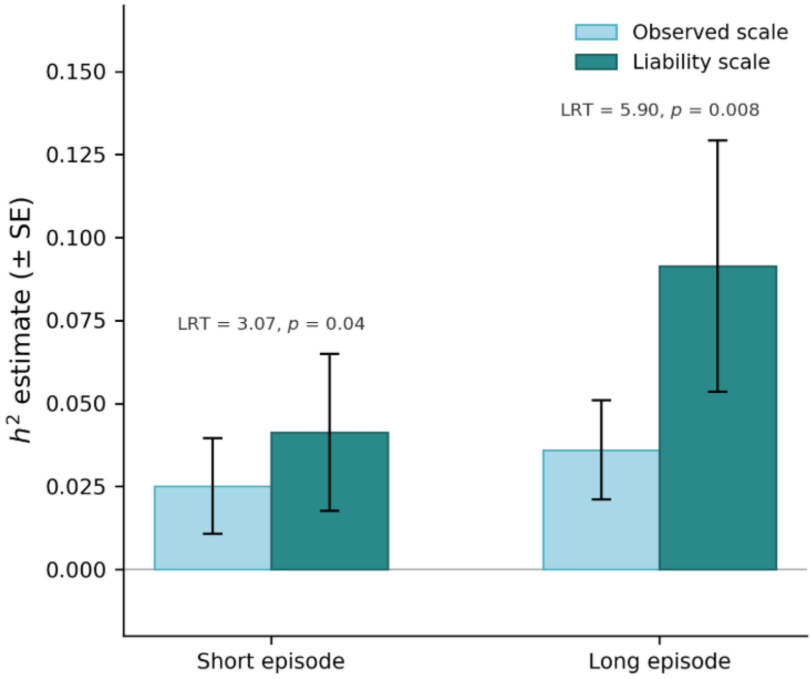


**Figure S5. SNP-based heritability of MDD episode duration.** Observed-scale (light blue) and liability-scale (dark teal) $h^2$ estimates (± SE) for long and short episode duration, estimated using GREML in unrelated EUR-like participants.

### Section 9: Full logistic regression results

**Table S6.** Full logistic regression results for short episode duration (0-3 months), showing odds ratios, 95% confidence intervals, and raw and Bonferroni-adjusted p-values for each predictor across the univariable, domain-specific, and combined models.

| **Predictor** | **Univariable** | | | | **Domain-specific** | | | | **Combined** | | |
|---|---|---|---|---|---|---|---|---|---|---|---|
| **Genetic variables (PGS)** | **N** | **OR [95% CI]** | ***p*** | ***p**bon* | **N** | **OR [95% CI]** | ***p*** | ***p**bon* | **N** | **OR [95% CI]** | ***p*** |
| MDD | 33013 | 0.904 [0.882 - 0.926] | < 0.001 | **< 0.001** | 33013 | 0.912 [0.888 - 0.938] | < 0.001 | **< 0.001** | 11710 | 0.943 [0.899 - 0.988] | **0.015** |
| ADHD | 33013 | 0.949 [0.928 - 0.972] | < 0.001 | **< 0.001** | 33013 | 0.980 [0.956 - 1.004] | 0.105 | 0.842 | 11710 | 0.983 [0.942 - 1.027] | 0.443 |
| Anorexia nervosa | 33013 | 1.031 [1.008 - 1.053] | 0.007 | 0.124 | 33013 | 1.017 [0.994 - 1.040] | 0.148 | 1.0 | 11710 | 1.021 [0.983 - 1.061] | 0.28 |
| Bipolar disorder | 33013 | 0.987 [0.964 - 1.010] | 0.258 | 1.0 | 33013 | 1.022 [0.996 - 1.048] | 0.097 | 0.777 | 11710 | 1.008 [0.965 - 1.054] | 0.715 |
| OCD | 33013 | 0.967 [0.946 - 0.989] | 0.003 | 0.063 | 33013 | 0.987 [0.964 - 1.010] | 0.272 | 1.0 | 11710 | 0.993 [0.954 - 1.034] | 0.74 |
| Schizophrenia | 33013 | 0.972 [0.949 - 0.996] | 0.023 | 0.444 | 33013 | 0.990 [0.964 - 1.016] | 0.437 | 1.0 | 11710 | 0.997 [0.953 - 1.043] | 0.899 |
| SSRI non-remission | 33013 | 0.976 [0.955 - 0.998] | 0.03 | 0.578 | 33013 | 0.983 [0.957 - 1.009] | 0.192 | 1.0 | 11710 | 1.007 [0.962 - 1.055] | 0.753 |
| SSRI % improvement | 33013 | 1.022 [0.999 - 1.047] | 0.063 | 1.0 | 33013 | 1.011 [0.983 - 1.040] | 0.444 | 1.0 | 11710 | 1.048 [0.998 - 1.100] | 0.058 |
| **Sociodemographic variables** | | | | | | | | | | | |
| Sex | NA | NA | NA | NA | 27251 | 0.734 [0.695 - 0.774] | < 0.001 | **< 0.001** | 11710 | 0.703 [0.645 - 0.766] | **< 0.001** |
| BMI | 32949 | 0.991 [0.987 - 0.996] | < 0.001 | **0.003** | 27251 | 0.993 [0.988 - 0.998] | 0.006 | **0.032** | 11710 | 1.000 [0.992 - 1.008] | 0.957 |
| Education | | | | | | | | | | | |
| None of the above | 32883 | 0.752 [0.674 - 0.838] | < 0.001 | **< 0.001** | 27251 | 0.797 [0.705 - 0.901] | < 0.001 | **0.001** | 11710 | 1.002 [0.811 - 1.239] | 0.982 |
| Secondary | 32883 | 0.851 [0.803 - 0.901] | < 0.001 | **< 0.001** | 27251 | 0.870 [0.816 - 0.927] | < 0.001 | **< 0.001** | 11710 | 0.904 [0.816 - 1.001] | 0.052 |
| Further | 32883 | 0.897 [0.838 - 0.959] | 0.002 | **0.03** | 27251 | 0.889 [0.825 - 0.958] | 0.002 | **0.01** | 11710 | 0.879 [0.783 - 0.988] | **0.03** |
| Vocational | 32883 | 0.922 [0.856 - 0.993] | 0.033 | 0.619 | 27251 | 0.939 [0.865 - 1.020] | 0.139 | 0.694 | 11710 | 0.931 [0.813 - 1.066] | 0.299 |
| TDI | 32972 | 0.980 [0.972 - 0.988] | < 0.001 | **< 0.001** | 27251 | 0.985 [0.976 - 0.994] | < 0.001 | **0.004** | 11710 | 0.982 [0.968 - 0.997] | **0.015** |
| Neuroticism score | 27428 | 0.939 [0.932 - 0.946] | < 0.001 | **< 0.001** | 27251 | 0.941 [0.934 - 0.948] | < 0.001 | **< 0.001** | 11710 | 0.977 [0.964 - 0.990] | **< 0.001** |
| **Clinical variables** | | | | | | | | | | | |
| Number of episodes | | | | | | | | | | | |
| 2-3 | 30556 | 0.806 [0.756 - 0.860] | < 0.001 | **< 0.001** | 13647 | 0.755 [0.676 - 0.843] | < 0.001 | **< 0.001** | 11710 | 0.750 [0.666 - 0.845] | **< 0.001** |
| 4-5 | 30556 | 0.828 [0.766 - 0.896] | < 0.001 | **< 0.001** | 13647 | 0.758 [0.656 - 0.875] | < 0.001 | **0.001** | 11710 | 0.757 [0.647 - 0.885] | **< 0.001** |
| 6+ | 30556 | 0.808 [0.733 - 0.890] | < 0.001 | **< 0.001** | 13647 | 0.752 [0.631 - 0.896] | 0.001 | **0.01** | 11710 | 0.757 [0.624 - 0.918] | **0.005** |

| | | | | | | | | | | | |
|---|---|---|---|---|---|---|---|---|---|---|---|
| Too many to count | 30556 | 0.339 [0.317 - 0.362] | < 0.001 | **< 0.001** | 13647 | 0.279 [0.240 - 0.323] | < 0.001 | **< 0.001** | 11710 | 0.294 [0.250 - 0.345] | **< 0.001** |
| Age of onset | 29201 | 1.003 [1.002 - 1.005] | < 0.001 | **< 0.001** | 13647 | 0.995 [0.992 - 0.998] | 0.002 | **0.014** | 11710 | 0.993 [0.990 - 0.997] | **< 0.001** |
| Years since last episode | 28279 | 1.014 [1.012 - 1.016] | < 0.001 | **< 0.001** | 13647 | 1.001 [0.997 - 1.005] | 0.737 | 1.0 | 11710 | 1.000 [0.995 - 1.004] | 0.855 |
| Stressful trigger | 32905 | 0.830 [0.790 - 0.873] | < 0.001 | **< 0.001** | 13647 | 0.757 [0.696 - 0.822] | < 0.001 | **< 0.001** | 11710 | 0.752 [0.687 - 0.823] | **< 0.001** |
| Childhood trauma score | 21710 | 0.893 [0.872 - 0.915] | < 0.001 | **< 0.001** | 13647 | 0.939 [0.909 - 0.969] | < 0.001 | **< 0.001** | 11710 | 0.948 [0.916 - 0.981] | **0.002** |
| Family history | 32684 | 0.807 [0.771 - 0.846] | < 0.001 | **< 0.001** | 13647 | 0.866 [0.803 - 0.933] | < 0.001 | **0.001** | 11710 | 0.894 [0.824 - 0.970] | **0.007** |
| Depression subtype | | | | | | | | | | | |
| Increased weight/sleep | 26486 | 0.668 [0.596 - 0.749] | < 0.001 | **< 0.001** | 13647 | 0.762 [0.641 - 0.905] | 0.002 | **0.014** | 11710 | 0.784 [0.651 - 0.944] | **0.01** |
| Decreased weight/sleep | 26486 | 0.917 [0.870 - 0.968] | 0.002 | **0.029** | 13647 | 0.901 [0.835 - 0.972] | 0.007 | 0.051 | 11710 | 0.894 [0.823 - 0.972] | **0.008** |

ADHD: attention-deficit/hyperactivity disorder; BMI: body mass index; CI: confidence interval; MDD: major depressive disorder; OCD: obsessive compulsive disorder; OR: odds ratio; PGS: polygenic scores; SSRI: selective serotonin reuptake inhibitor; TDI: Townsend Deprivation Index.

**Table S7.** Full logistic regression results for long episode duration (>24 months), showing odds ratios, 95% confidence intervals, and raw and Bonferroni-adjusted p-values for each predictor across the univariable, domain-specific, and combined models.

| Predictor | Univariable | | | | Domain-specific | | | | Combined | | |
|---|---|---|---|---|---|---|---|---|---|---|---|
| **Genetic variables (PGS)** | **N** | **OR [95% CI]** | ***p*** | $p_{bon}$ | **N** | **OR [95% CI]** | ***p*** | $p_{bon}$ | **N** | **OR [95% CI]** | ***p*** |
| MDD | 33013 | 1.165 [1.125 - 1.206] | < 0.001 | **< 0.001** | 33013 | 1.119 [1.076 - 1.163] | < 0.001 | **< 0.001** | 11710 | 0.986 [0.915 - 1.062] | 0.702 |
| ADHD | 33013 | 1.100 [1.064 - 1.137] | < 0.001 | **< 0.001** | 33013 | 1.047 [1.011 - 1.085] | 0.01 | 0.079 | 11710 | 1.026 [0.958 - 1.098] | 0.464 |
| Anorexia nervosa | 33013 | 0.952 [0.924 - 0.982] | 0.002 | **0.036** | 33013 | 0.980 [0.950 - 1.012] | 0.214 | 1.0 | 11710 | 0.948 [0.893 - 1.006] | 0.08 |
| Bipolar disorder | 33013 | 1.058 [1.023 - 1.093] | < 0.001 | **0.017** | 33013 | 0.994 [0.958 - 1.031] | 0.738 | 1.0 | 11710 | 1.040 [0.971 - 1.115] | 0.263 |
| OCD | 33013 | 1.084 [1.050 - 1.119] | < 0.001 | **< 0.001** | 33013 | 1.047 [1.013 - 1.082] | 0.006 | 0.051 | 11710 | 1.009 [0.947 - 1.076] | 0.778 |
| Schizophrenia | 33013 | 1.091 [1.054 - 1.129] | < 0.001 | **< 0.001** | 33013 | 1.053 [1.014 - 1.093] | 0.007 | 0.054 | 11710 | 1.026 [0.956 - 1.101] | 0.481 |
| SSRI non-remission | 33013 | 1.026 [0.994 - 1.058] | 0.109 | 1.0 | 33013 | 1.026 [0.988 - 1.065] | 0.179 | 1.0 | 11710 | 1.013 [0.943 - 1.089] | 0.72 |
| SSRI % improvement | 33013 | 0.988 [0.955 - 1.021] | 0.462 | 1.0 | 33013 | 1.004 [0.965 - 1.045] | 0.839 | 1.0 | 11710 | 0.978 [0.906 - 1.056] | 0.573 |
| **Sociodemographic variables** | | | | | | | | | | | |
| Sex | NA | NA | NA | NA | 27251 | 0.914 [0.845 - 0.988] | 0.023 | 0.117 | 11710 | 1.019 [0.888 - 1.168] | 0.793 |
| BMI | 32949 | 1.024 [1.017 - 1.030] | < 0.001 | **< 0.001** | 27251 | 1.019 [1.012 - 1.026] | < 0.001 | **< 0.001** | 11710 | 1.001 [0.989 - 1.013] | 0.839 |
| Education | | | | | | | | | | | |
| None of the above | 32883 | 1.793 [1.566 - 2.052] | < 0.001 | **< 0.001** | 27251 | 1.454 [1.244 - 1.699] | < 0.001 | **< 0.001** | 11710 | 1.191 [0.881 - 1.610] | 0.256 |
| Secondary | 32883 | 1.378 [1.271 - 1.495] | < 0.001 | **< 0.001** | 27251 | 1.294 [1.181 - 1.417] | < 0.001 | **< 0.001** | 11710 | 1.200 [1.024 - 1.407] | **0.024** |
| Further | 32883 | 1.201 [1.089 - 1.324] | < 0.001 | **0.004** | 27251 | 1.087 [0.972 - 1.214] | 0.143 | 0.715 | 11710 | 1.213 [1.013 - 1.452] | **0.036** |
| Vocational | 32883 | 1.337 [1.204 - 1.484] | < 0.001 | **< 0.001** | 27251 | 1.228 [1.091 - 1.383] | < 0.001 | **0.003** | 11710 | 1.260 [1.028 - 1.546] | **0.026** |
| TDI | 32972 | 1.045 [1.034 - 1.056] | < 0.001 | **< 0.001** | 27251 | 1.041 [1.028 - 1.053] | < 0.001 | **< 0.001** | 11710 | 1.020 [0.998 - 1.042] | 0.075 |
| Neuroticism score | 27428 | 1.148 [1.135 - 1.161] | < 0.001 | **< 0.001** | 27251 | 1.143 [1.130 - 1.156] | < 0.001 | **< 0.001** | 11710 | 1.053 [1.033 - 1.075] | **< 0.001** |
| **Clinical variables** | | | | | | | | | | | |
| Number of episodes | | | | | | | | | | | |
| 2-3 | 30556 | 1.211 [1.064 - 1.379] | 0.004 | 0.07 | 13647 | 1.304 [1.067 - 1.595] | 0.01 | 0.067 | 11710 | 1.314 [1.054 - 1.639] | **0.015** |
| 4-5 | 30556 | 1.156 [0.987 - 1.353] | 0.072 | 1.0 | 13647 | 1.081 [0.834 - 1.400] | 0.556 | 1.0 | 11710 | 1.066 [0.801 - 1.417] | 0.662 |
| 6+ | 30556 | 1.609 [1.353 - 1.913] | < 0.001 | **< 0.001** | 13647 | 1.371 [1.023 - 1.836] | 0.035 | 0.243 | 11710 | 1.449 [1.055 - 1.990] | **0.022** |
| Too many to count | 30556 | 7.564 [6.869 - 8.330] | < 0.001 | **< 0.001** | 13647 | 6.715 [5.515 - 8.177] | < 0.001 | **< 0.001** | 11710 | 6.127 [4.925 - 7.623] | **< 0.001** |

| | | | | | | | | | | | |
|---|---|---|---|---|---|---|---|---|---|---|---|
| Age of onset | 29201 | 0.982 [0.980 - 0.984] | < 0.001 | **< 0.001** | 13647 | 0.996 [0.992 - 1.000] | 0.056 | 0.393 | 11710 | 0.998 [0.993 - 1.002] | 0.329 |
| Years since last episode | 28279 | 0.967 [0.964 - 0.970] | < 0.001 | **< 0.001** | 13647 | 0.990 [0.984 - 0.996] | < 0.001 | **0.006** | 11710 | 0.990 [0.984 - 0.997] | **0.005** |
| Stressful trigger | 32905 | 0.899 [0.837 - 0.964] | 0.003 | 0.056 | 13647 | 1.193 [1.050 - 1.356] | 0.007 | **0.047** | 11710 | 1.182 [1.028 - 1.360] | **0.019** |
| Childhood trauma score | 21710 | 1.276 [1.239 - 1.315] | < 0.001 | **< 0.001** | 13647 | 1.126 [1.079 - 1.175] | < 0.001 | **< 0.001** | 11710 | 1.127 [1.075 - 1.181] | **< 0.001** |
| Family history | 32684 | 1.310 [1.227 - 1.398] | < 0.001 | **< 0.001** | 13647 | 0.972 [0.865 - 1.092] | 0.629 | 1.0 | 11710 | 0.989 [0.871 - 1.125] | 0.871 |
| Depression subtype | | | | | | | | | | | |
| Increased weight/sleep | 26486 | 1.543 [1.356 - 1.755] | < 0.001 | **< 0.001** | 13647 | 1.074 [0.869 - 1.326] | 0.51 | 1.0 | 11710 | 1.031 [0.816 - 1.302] | 0.798 |
| Decreased weight/sleep | 26486 | 0.751 [0.695 - 0.811] | < 0.001 | **< 0.001** | 13647 | 0.872 [0.771 - 0.987] | 0.03 | 0.208 | 11710 | 0.833 [0.727 - 0.955] | **0.009** |

ADHD: attention-deficit/hyperactivity disorder; BMI: body mass index; CI: confidence interval; MDD: major depressive disorder; OCD: obsessive compulsive disorder; OR: odds ratio; PGS: polygenic scores; SSRI: selective serotonin reuptake inhibitor; TDI: Townsend Deprivation Index.
